# A Culturally Embedded Augmented Reality Task as a Neurocognitive Biomarker of Executive Function in Schizophrenia

**DOI:** 10.64898/2026.07.14.26358053

**Authors:** W. Chatthong, M. Rueankam, S. Khemthong

## Abstract

**Background:** Executive function (EF) deficits are central features of schizophrenia and strongly influence long-term functional outcomes. Conventional cognitive assessments often lack ecological validity and cultural relevance.

**Aim:** This study introduces the Luk Chup Augmented Reality (LCAR) tool—a video-guided, clay modeling task delivered through wearable AR—that integrates culturally familiar activity with real-time neurophysiological monitoring.

**Methods:** Thirty individuals diagnosed with schizophrenia (mean age = 38.9 ± 7.15 years) completed a series of modeling and memory tasks using LCAR while undergoing quantitative EEG (QEEG). Task duration and theta/beta power were analyzed across procedural and color-shape memory phases.

**Results:** Memory phases took significantly longer to complete and were associated with decreased lateral prefrontal theta and increased frontal-midline theta activity (Fz, Cz), indicating higher EF demand. A repeated-measures ANOVA revealed significant condition, site, and interaction effects on theta power.

**Conclusion:** The LCAR tool shows promise as a culturally grounded, dual-mode assessment of EF in schizophrenia. It offers a novel integration of performance-based and neurophysiological metrics that may inform future interventions in psychiatric rehabilitation.

**Graphic Abstract:** 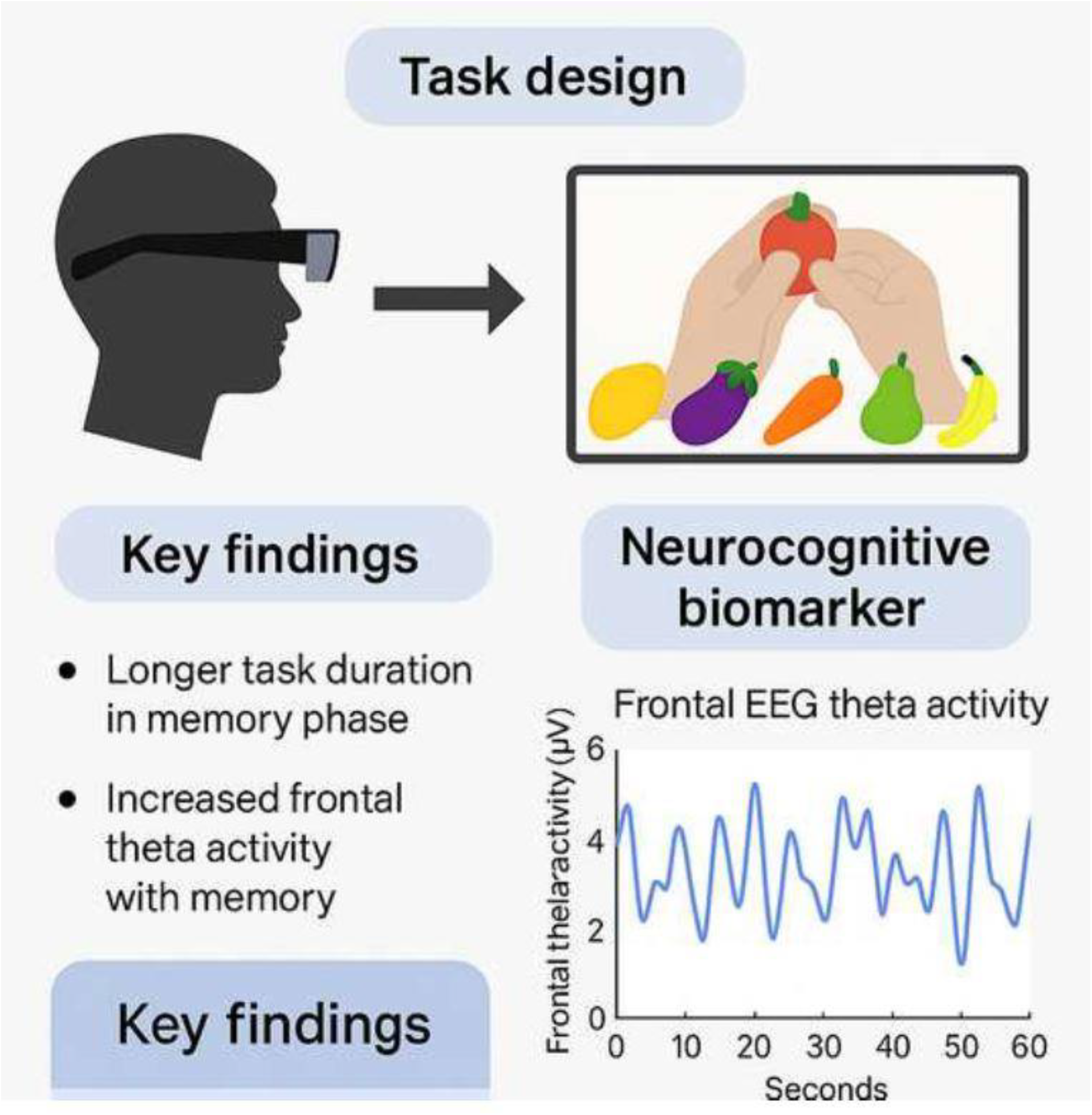

## 1. Introduction

Schizophrenia is a severe and heterogeneous neuropsychiatric disorder marked by persistent disruptions in cognitive domains, with executive function (EF) impairments emerging as central predictors of long-term functional disability (Kluwe-Schiavon et al., 2013; Cavanagh and Frank, 2014). Deficits in EF—encompassing planning, working memory, inhibition, and cognitive flexibility—limit individuals’ capacity to initiate and complete purposeful, goal-directed activities, directly impacting social participation and independent living (Tang et al., 2019; Synovec, 2015).

Conventional neuropsychological assessments often fall short in ecological validity; they isolate cognitive functions in decontextualized test environments, which may inadequately represent real-world functional performance (Giglioli et al., 2019; Sklar et al., 2020). This limitation has catalyzed interest in embedded, task-based tools that simulate naturalistic activity while capturing cognitive load (Leenerts et al., 2016; Khemthong and Wee, 2016).

Augmented reality (AR) technologies offer promising platforms for cognitive assessment by enabling real-time, interactive simulations of everyday tasks (Giglioli et al., 2019; Nahum et al., 2021). In the Thai context, the traditional fruit-shaped clay art known as *Luk Chup* has been culturally adapted into a cognitively engaging, AR-based modeling task—the Luk Chup Augmented Reality (LCAR) tool. LCAR merges culturally familiar craft with guided visual prompts and memory trials that invoke multiple EF components (Khemthong and Schrepp, 2019; Hinz, 2020). Critically, it is embedded in a format that promotes client engagement and minimizes task alienation (Su et al., 2011).

This study investigates the potential of LCAR as a performance-based, culturally grounded biomarker for EF in individuals with schizophrenia.

Developed through participatory input from service users and iterative usability testing, the tool is designed to be both neurologically informative and behaviorally relevant (Choi et al., 2020). By integrating task performance metrics with quantitative EEG (QEEG) data, we examine LCAR’s capacity to reflect neural dynamics associated with executive processing and its promise as a sensitive, contextually meaningful cognitive assessment in psychiatric rehabilitation.

## 2. Methodology

### 2.1. Participants

Thirty individuals (mean age = 38.9 ± 7.15 years; 10 male-identifying, 20 female-identifying; all right-handed), diagnosed with schizophrenia, were recruited from outpatient psychiatric services at the Psychosocial Occupational Therapy Clinic, Faculty of Physical Therapy, Mahidol University. All participants met DSM-5 diagnostic criteria and provided written informed consent. To ensure clinical stability, participants were screened using the Thai version of the Positive and Negative Syndrome Scale (PANSS) prior to electroencephalographic (EEG) recording. Individuals with acute psychotic symptoms or high positive symptom scores were excluded from testing. The PANSS Thai version has demonstrated acceptable criterion validity and interrater reliability (Nilchaikovit et al., 2000). Additional exclusion criteria included neurological disorders and current substance use.

### 2.2. Ethical Approval

The study was approved by the Central Institutional Review Board of Mahidol University (MU-CIRB Project ID: 2020/114.1505; COA No. 2020/103.0708) and the Ethics Committee of the Department of Mental Health, Thailand (DMH.IRB 031/2563; COA No. 041/2563).

### 2.3. LCAR Task Creation

The Luk Chup Augmented Reality (LCAR) tool was developed as a novel, culturally grounded cognitive task based on the Thai fruit-shaped clay art known as *Luk Chup*. The tool employs a Video-based Augmented Reality (VAR) interface delivered through a wearable optical display. LCAR was designed to activate frontal and temporal regions involved in object recognition, visuospatial modeling, and working memory through guided video instructions and hands-on clay manipulation. Task elements were informed by Piagetian developmental theory, occupational therapy craft analysis (Hinz, 2020; Leenerts et al., 2016), and tactile memory research (Ayres, 1985).

User interface design was iteratively refined based on feedback from 10 volunteers with mental health experience, who also completed the Thai version of the User Experience Questionnaire (UEQ v.7), demonstrating above-threshold clarity (1.93) and effectiveness (1.64) scores (Khemthong and Schrepp, 2019).

The task sequence was aligned with Allen Cognitive Levels 4–5 (Su et al., 2011), focusing on goal-directed behavior, visual discrimination, and problem-solving. The LCAR protocol consists of seven stages that increase in complexity, requiring participants to complete six clay fruit models—yellow mango, purple eggplant, orange carrot, green pear, yellow-green papaya, and red rose apple— followed by a color-shape memory task. Completion time was recorded for each stage. Augmented and extended reality platforms have increasingly been recognized for their potential in cognitive assessment, offering ecological validity and immersive engagement (Bello et al., 2025) (see Figure 1).

**Fig. 1.**
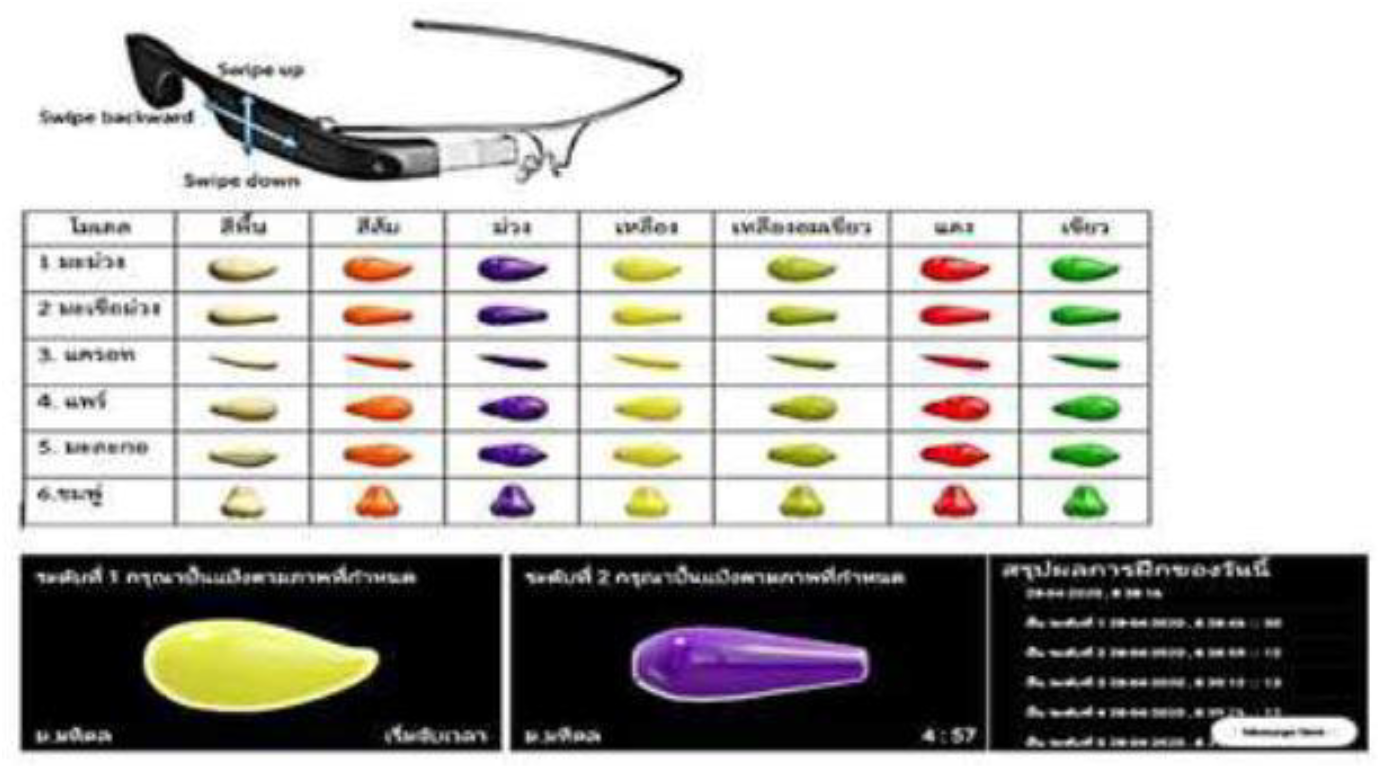
Structure of the Luk Chup Augmented Reality (LCAR) task, showing sequential stages of clay modeling and the final color-shape memory trial delivered through a wearable augmented reality interface.

### 2.4. Electrophysiological Analysis

Quantitative EEG (QEEG) was recorded throughout the LCAR protocol using a 12-channel system following the international 10–20 montage. Primary regions of interest included Fp1, Fz, F4, F7, Cz, and T3. Relative power in the theta band (4–7 Hz) was analyzed across three phases: baseline rest (eyes open), procedural modeling, and color-shape memory. A repeated-measures ANOVA was conducted to assess within-subject differences in theta activity across task conditions and recording sites.

## 3. Results

Task completion times increased with cognitive complexity. The mean durations (mean ± SD) for the six LCAR crafting tasks ranged from 45.26 ± 26.30 seconds to 61.00 ± 42.41 seconds, while the final color-shape memorization task required significantly more time, with a mean of 142.09 ± 115.25 seconds (Table 1).

**Table 1.** Mean completion time (sec) for each LCAR phase.

| LCAR Phase | Time (sec) ± SD |
| --- | --- |
| Craft 1: Yellow mango | $52.83 \pm 33.10$ |
| Craft 2: Purple eggplant | $57.78 \pm 37.57$ |
| Craft 3: Orange carrot | $51.61 \pm 27.07$ |
| Craft 4: Green pear | $45.26 \pm 26.30$ |
| Craft 5: Yellow-green papaya | $61.00 \pm 42.41$ |
| Craft 6: Red rose apple | $57.39 \pm 48.47$ |
| Color-shape memory | $142.09 \pm 115.25$ |

Theta relative power decreased across LCAR phases compared to resting states, particularly at lateral prefrontal sites (Fp1, T3). The lowest theta values were observed during the color-shape memory phase, suggesting increased cognitive and attentional demand. In contrast, frontal-midline regions (Fz, Cz) maintained relatively higher theta power throughout the task sequence, peaking during memory phases and reflecting sustained executive engagement (Table 2).

**Table 2.** Mean theta relative power (µV^2^) across QEEG sites and task phases.

| Site | Calm (Eyes Closed) | Rest (Eyes Open) | Crafting Phase | Color Memory |
| --- | --- | --- | --- | --- |
| Fp1 | 20.50 ± 9.15 | 16.51 ± 4.97 | 13.86 ± 6.23 | 11.89 ± 4.50 |
| F7 | 19.61 ± 8.42 | 18.45 ± 5.04 | 16.27 ± 5.45 | 15.12 ± 2.16 |
| F4 | 21.97 ± 10.23 | 20.75 ± 5.47 | 18.43 ± 6.01 | 17.70 ± 5.04 |
| Fz | 23.87 ± 10.88 | 22.99 ± 6.42 | 21.31 ± 7.30 | 21.99 ± 4.63 |
| Cz | 22.52 ± 11.76 | 21.95 ± 6.20 | 20.50 ± 5.34 | 21.16 ± 4.48 |
| T3 | 17.37 ± 9.50 | 16.29 ± 6.83 | 14.11 ± 6.25 | 12.65 ± 5.22 |

This sequential progression from guided crafting to independent color-shape recall mirrors the design of the LCAR task itself, in which increasingly complex cognitive demands are embedded across stages. As shown in Figure 2, theta activity declined across lateral prefrontal sites (Fp1, T3) during this progression, while frontal-midline theta (Fz, Cz) remained elevated—consistent with sustained executive control.

**Fig. 2.**
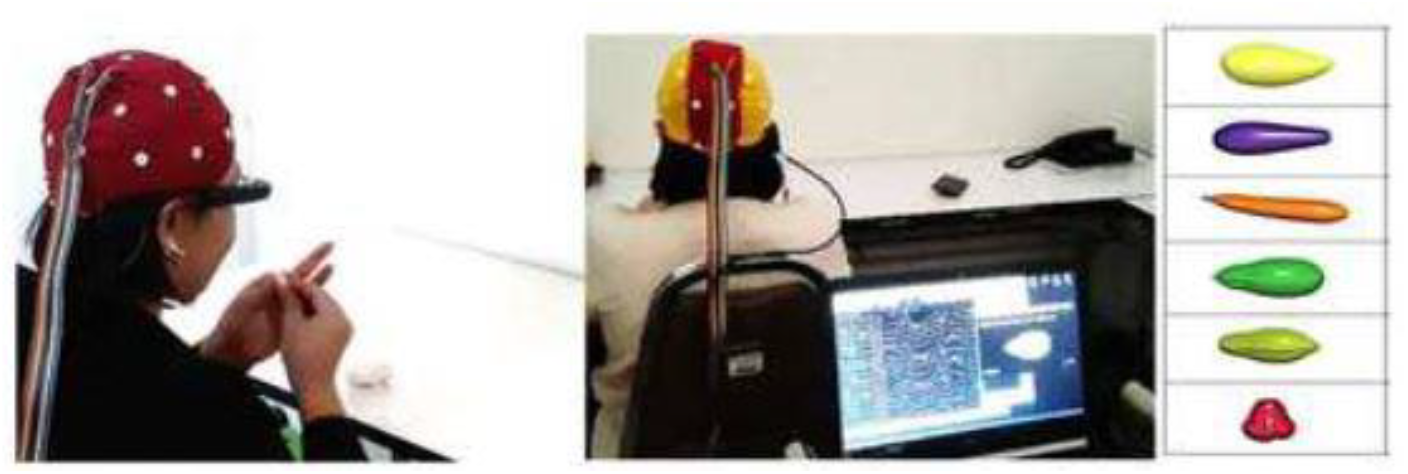
Relative theta power across six QEEG sites (Fp1, F7, F4, Fz, Cz, T3) during four LCAR phases: calm (eyes closed), rest (eyes open), crafting, and color memory. The pattern reflects cognitive load intensification aligned with LCAR task design, progressing from modeling to explicit memory retrieval.

Theta activity progressively declined from calm to rest to LCAR performance. The most pronounced reductions were observed at Fp1 and T3 during the color memory phase, consistent with increased executive and attentional demands. In contrast, Fz and Cz exhibited consistently elevated theta levels, reflecting stable executive engagement across all cognitive conditions. Frontal-midline theta (especially Fz) remained highest across all conditions (peak =23.87 ± 10.88 during calm; lowest = 21.31 ± 7.30 during crafting), supporting its role in attention and cognitive control.

A repeated-measures ANOVA (4 conditions × 6 electrode sites) revealed a significant main effect of condition, F(1.648, 36.258) = 4.264, p = .028; a significant main effect of site, F(1.868, 41.100) = 31.271, p < .001; and a condition-by-site interaction, F(6.643, 146.146) = 5.220, p < .001. These findings confirm region-specific modulation of theta power in response to executive demands, particularly during the transition from procedural to memory-based tasks. This dynamic aligns with recent work showing anterior prefrontal theta fluctuations as markers of working memory and executive functioning (Hamedi, et al., 2025).

## 4. Discussion

The Luk Chup Augmented Reality (LCAR) protocol engaged multiple facets of executive functioning through a culturally meaningful, interactive format. The prolonged completion time observed during the color-shape memory phase, coupled with sustained frontal-midline theta activity at Fz and Cz, reflects heightened executive demands—particularly in the domains of working memory, set-shifting, and attentional control (Cavanagh and Frank, 2014; Giglioli et al., 2019; Tang et al., 2019). Repeated-measures ANOVA confirmed a significant interaction between condition and site, with Fz and Cz consistently showing the highest theta power across task stages, reinforcing their role in sustained cognitive engagement.

The extended task duration during memory phases may represent inhibition latency or transient overload in executive processing, consistent with dual-task interference and set-shifting impairments commonly observed in schizophrenia (Kluwe-Schiavon et al., 2013; Synovec, 2015). These patterns align with existing cognitive remediation strategies that employ graduated task complexity—such as multi-step activities like grooming or meal preparation—to train flexibility, working memory, and attentional redirection (Leenerts et al., 2016).

Within this conceptual framework, LCAR may serve not only as a performance-based EF assessment but also as a therapeutic tool within longitudinal cognitive rehabilitation programs. Its structure mirrors emerging Thai executive rehabilitation models that emphasize stepwise complexity, real-time cueing, and affective readiness to enhance engagement and reduce task alienation (Khemthong and Wee, 2016).

This study has several limitations. First, the sample size was modest and cross-sectional, which may limit generalizability and preclude conclusions about longitudinal change. Second, the study did not include a healthy control group, restricting comparative interpretation of QEEG patterns. Third, while the LCAR tool was designed with cultural relevance in mind, its applicability to individuals from other cultural or linguistic backgrounds remains to be tested. Lastly, the neurophysiological data focused primarily on theta activity; future studies could benefit from including additional frequency bands and connectivity analyses.

From a translational perspective, culturally embedded tools like LCAR can improve the sensitivity of cognitive screening while fostering psychological safety and intrinsic motivation. This is particularly important in mental health contexts where traditional assessments may be perceived as foreign or disconnected from daily life (Synovec, 2015; Nahum et al., 2021; Waanicharuedi et al., 2020). Recent developments in augmented reality–based therapy for negative symptoms in schizophrenia further support the feasibility of such tools in psychiatric rehabilitation (Tang et al., 2024).

The familiarity and structure of LCAR may also support therapeutic rapport by reducing performance anxiety and promoting user confidence. Neurophysiologically, the observed increases in frontal-midline theta and task-specific beta modulation support the use of LCAR as a dual-mode biomarker, integrating behavioral performance with region-specific neural signatures (Giglioli et al., 2019; Su et al., 2011). This alignment between task complexity and QEEG indicators strengthens the tool’s potential utility in real-time cognitive monitoring and tailored intervention planning.

Future research should examine LCAR’s test–retest reliability, sensitivity to change over time, and predictive value in broader clinical contexts. Investigating its integration into digital cognitive training systems or community-based rehabilitation platforms may further advance its impact on person-centered care in schizophrenia and related conditions.

## 5. Conclusion

The Luk Chup Augmented Reality (LCAR) task represents a promising, culturally grounded approach to executive function assessment in schizophrenia. By integrating performance-based metrics with neurophysiological markers in real time, LCAR provides a dual-mode framework that enhances both diagnostic precision and intervention planning. Its immersive, familiar format may increase ecological validity, client engagement, and translational applicability in cognitive rehabilitation. Future implementation in longitudinal and community-based settings may further support its role as a practical tool for personalized mental health care.

## CRediT authorship contribution statement

W.C.: Writing – review & editing, Supervision, Methodology, Investigation, Data curation. M. R.: Writing – review & editing, Data curation. S. K.: Writing – original draft, Investigation, Formal analysis, Data curation, Conceptualization, Funding acquisition, Project administration.

## Declaration of generative AI and AI-assisted technologies in the writing process

During the preparation of this work the author(s) used ChatGPT (OpenAI, 2024) to improve clarity, ensure consistency in formatting, and support scientific writing refinement. After using this tool, the author(s) reviewed and edited the content as needed and take full responsibility for the content of the published article.

## Data Availability

All data produced in the present study are available upon reasonable request to the authors.

## Acknowledgments

The authors sincerely thank the participants and mental health professionals who contributed their time and insights to this study.

